# Itopride hydrochloride extended-release vs film-coated tablets for gastrointestinal symptoms in functional dyspepsia

**DOI:** 10.64898/2026.07.31.26359023

**Authors:** Sakkarin Chirapongsathorn, Marc Alison P. Hizon, Sanjiv Mahadeva, Anush Sargsyan, Nguyen Cong Long, Nguyen Thi Nha Doan, Pham Quang Phu, Suntje Sander

**Affiliations:** Phramongkutklao Hospital and Phramongkutklao College of Medicine, Bangkok, Thailand; San Fernando Interchange, Pampanga, Philippines; Department of Medicine, Faculty of Medicine, University Malaya, Kuala Lumpur, Malaysia; Grigor Narekatsi Medical Center, Yerevan, Armenia; Bach Mai Hospital (BMH), Hanoi, Vietnam; Nguyen Tri Phuong Hospital, Ho Chi Minh City, Vietnam; Military Hospital 103, Hanoi, Vietnam; Abbott Laboratories GmbH, Hannover, Germany

## Abstract

**Background:** Functional dyspepsia (FD) is among the most common gastrointestinal disorders worldwide and is characterized by symptoms including epigastric pain, early satiety, postprandial fullness, bloating, and upper abdominal discomfort. Itopride hydrochloride is commonly administered as 50 mg three times daily (TID). To improve convenience and potentially enhance adherence, a once-daily (OD) 150 mg extended-release formulation was developed. Phase 1 studies demonstrated bioequivalent overall exposure between the OD and TID regimens, with sustained-release characteristics and no evidence of dose dumping, supporting advancement to Phase 3. This pivotal clinical study evaluated whether itopride hydrochloride 150 mg OD is non-inferior to the established 50 mg TID regimen in improving FD symptoms over 8 weeks.

**Methods:** This Phase 3, randomized, open-label, multicenter, active-controlled study enrolled 564 participants with FD (or chronic gastritis) to compare the efficacy and safety of itopride hydrochloride 150 mg OD versus 50 mg TID over 8 weeks. The primary endpoint was change in overall FD severity from baseline to Week 8, assessed using the Leeds Dyspepsia Questionnaire (LDQ) severity score. Secondary endpoints included symptom-specific severity, disease-specific quality of life, responder rates, treatment acceptance, and safety.

**Results:** Clinical non-inferiority in terms of overall FD severity was demonstrated with OD treatment compared to TID. LDQ severity improved by −9.60 (95% CI −10.15, −9.05) with TID and −9.76 (95% CI −10.32, −9.19) with OD, with a between-group difference of 0.16 (95% CI −0.42, 0.74). Improvements across symptom domains and quality of life measures were comparable. Treatment acceptance favored OD (mean 4.22 vs 3.83; p < 0.001). Both regimens were well tolerated, with predominantly mild adverse events. This clinical evaluation of OD was supported by the Phase 1 results confirming that both single-dose and multiple-dose administration of itopride hydrochloride 150 mg OD provided a comparable extent of exposure to the 50 mg TID regimen, demonstrated by area under the curve (AUC) values within the 80–125% bioequivalence range and stable pharmacokinetic profiles across fed and fasted conditions.

**Conclusion:** The study confirmed that itopride hydrochloride 150 mg OD is non-inferior to the TID regimen for improving FD (or chronic gastritis) symptoms. The OD regimen demonstrated comparable efficacy, favorable treatment acceptance, and a positive benefit-risk profile, offering a more convenient therapeutic option for patients.

**Clinical trials registration:** NCT06217393

## Introduction

Dyspepsia is defined as episodic and persistent symptoms of the upper gastrointestinal tract and is the medical term for difficult digestion. Dyspepsia is associated with various symptoms, including bloating, fullness, heartburn, early satiation, nausea, vomiting, and abdominal pain (1).

The global prevalence of dyspepsia is at least 20% but varies considerably between countries (from 1.8% to 57.0%). Dyspepsia prevalence is higher in women, smokers, non-steroidal anti-inflammatory drug users and Helicobacter pylori-positive individuals (2).

Based on the cause, dyspepsia is classified into two main categories, namely organic and Functional Dyspepsia (FD), otherwise known as gastric dyspepsia. Organic dyspepsia includes peptic ulcer and Gastroesophageal Reflux Disease (GERD) as the most identifiable causes. Whereas FD, which is one of the most prevalent Functional Gastrointestinal Disorders (FGIDs), is identified by pathophysiological mechanisms such as delayed gastric emptying, impaired gastric accommodation to a meal, burning sensation in the epigastrium, hypersensitivity to gastric distension, predominant epigastric pain, early satiety and feeling of fullness during or post-meals (3–5). Globally, most dyspepsia patients fall into the category of FD, also known as non-ulcer dyspepsia, and large population-based studies report an FD prevalence ranging from 10% to 30% worldwide (6). Patients with FD experience reduced health-related quality of life, with symptoms, such as abdominal pain, impacting mental health, physical and social functioning (7,8).

The diagnosis of FD can be challenging given the overlap with other symptoms, including upper abdominal pain or discomfort, nausea and vomiting, loss of appetite and bloating (9). Patients may have both conditions and gastritis can also be a factor in developing FD (10). FD is characterized by the continual presence of epigastric pain, bloating and nausea (10).

Treatments for FD include dietary modifications, acid suppressants, prokinetics, neuromodulators and behavioral therapies (11). Itopride hydrochloride, a prokinetic therapy with both dopamine D2 receptor antagonist and acetylcholinesterase inhibitor properties (12), is indicated for the treatment of gastrointestinal symptoms caused by gastric dysmotility and delayed gastric emptying, including the sensation of bloating, early satiety, postprandial fullness, upper abdominal pain or discomfort, anorexia, heartburn, nausea, and vomiting; functional (non-ulcer) dyspepsia or chronic gastritis. Of note, the clinical presentation and management of FD can vary between Asia and Europe/the United States owing to differences in healthcare practices, the prevalence of certain conditions and cultural factors. While the concept of FD was formally defined in Europe with the advent of the Rome criteria in the 1990s, in some Asian countries, FD has only recently been recognized as a distinct condition (13–16). Previously, only the diagnosis of chronic gastritis was used to define the combination of gastrointestinal symptoms. FD is now considered the modern terminology for chronic gastritis and is diagnosed using the Rome IV criteria, with a differentiation between postprandial fullness and epigastric pain syndrome (17). These developments have helped standardize the diagnosis and management of FD across different regions, taking into account cultural and physiological differences.

Itopride hydrochloride is commonly dosed at 50 mg three times a day (TID) for the treatment of FD. The benefit of reducing the dose from thrice-daily to once-daily (OD) that comes with the 150 mg extended-release OD preparation could therefore be a strategic life cycle management opportunity, with the reduction in frequency of dosing potentially helping to improve patient compliance and adherence. To date, 18 randomized controlled trials (six placebo-controlled and 12 reference-controlled trials), including a total of 4,410 patients, and two meta-analyses, each including approximately 2,500 patients, have provided evidence of the efficacy of a daily dose of 150 mg itopride hydrochloride for the treatment of FD.

This study aimed to investigate the non-inferiority of OD itopride hydrochloride 150 mg extended-release tablets versus TID itopride hydrochloride 50 mg film-coated tablets by examining efficacy and safety in participants with gastrointestinal symptoms caused by gastric dysmotility and delayed gastric emptying with FD (non-ulcer) or chronic gastritis.

## Methods

### Study design

From 28 February 2024 to 28 February 2025, this Phase 3, randomized, open-label, multicenter, parallel-group, active-controlled study compared an OD dose of 150 mg itopride hydrochloride extended-release tablet with a TID dose of 50 mg itopride hydrochloride film-coated tablets, in participants with gastrointestinal symptoms. These symptoms were a result of gastric dysmotility and delayed gastric emptying with FD (non-ulcer) or chronic gastritis and included bloating sensation, early satiety, postprandial fullness, upper abdominal pain or discomfort, anorexia, heartburn, nausea and vomiting. The study was conducted at 19 sites across 5 countries (Armenia, Malaysia, Philippines, Thailand, and Vietnam).

After screening, participants were randomized in a 1:1 ratio to itopride hydrochloride 150 mg extended-release tablets OD administered before one of the main meals (preferably the same meal throughout the treatment; OD group) or itopride hydrochloride 50 mg film-coated tablets TID administered before meals for 8 weeks (TID group). The study screened 769 participants of which 564 participants were enrolled (282 participants in each arm), and the duration of participation was approximately 11 weeks (**Figure 1**).

**Figure 1.**
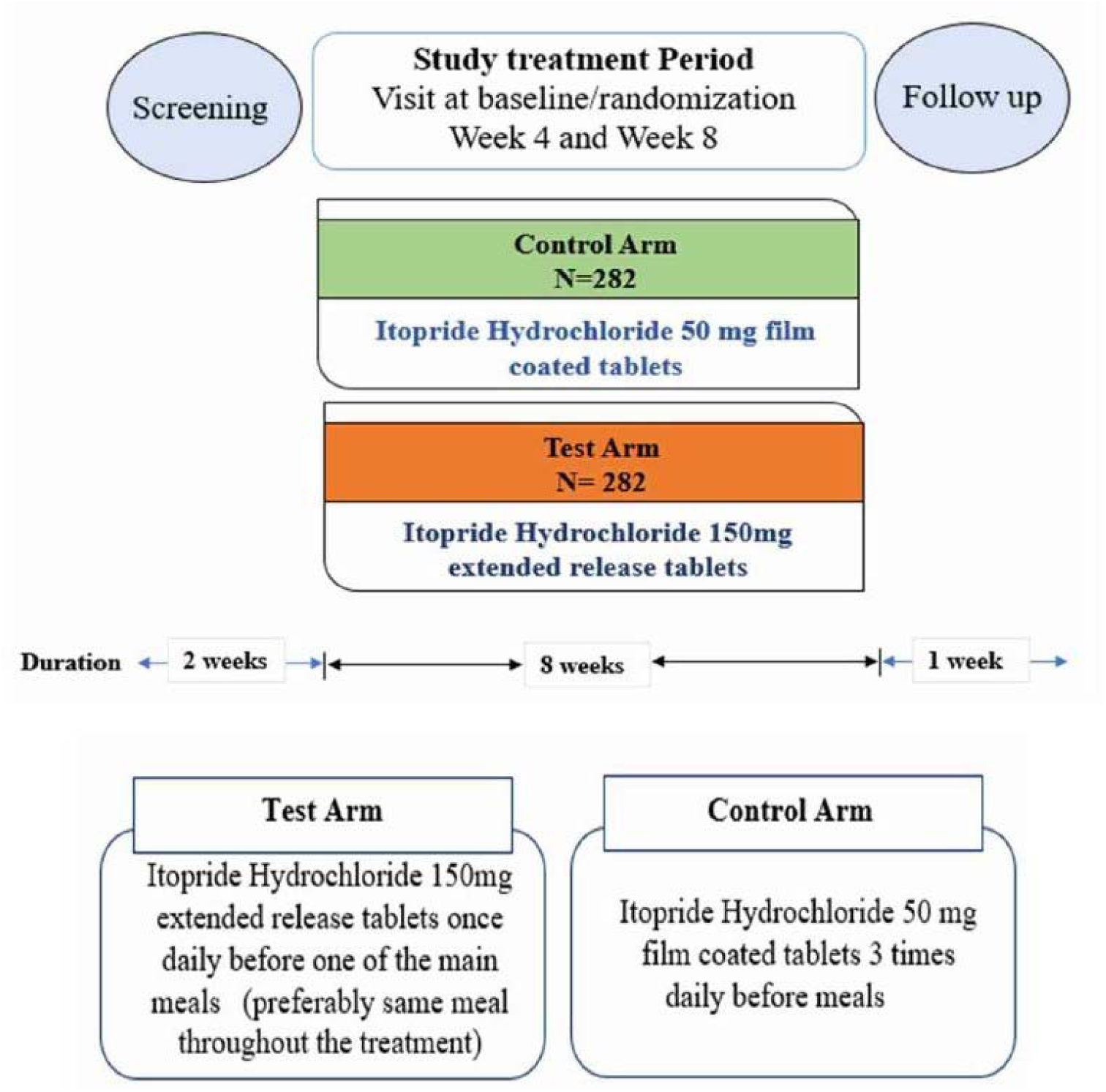
Phase 3 study Design

Participants were required to discontinue most gastric function related drugs prior to the study and participants taking acid-release inhibitors, antacids or gastric mucosa protectors were excluded. Rescue medications, including proton-pump inhibitors (PPIs) or antacids, for participants with persistent symptoms of gastric hypersecretion, were started after confirmation by the Investigator and not earlier than 4 weeks into the study (visit 3).

### Assessments

The primary endpoint of the phase III study was the change in overall severity of FD from baseline to Week 8, as measured by the LDQ severity score. Secondary endpoints included the change in overall FD severity between baseline and Week 4 assessed by the LDQ severity score, Disease Specific Quality of Life (QoL) assessed by the Short Form Nepean Dyspepsia Index (SF-NDI) at baseline and Week 8, change from baseline in the Numerical Rating Scale – 11 (NRS-11) score for symptoms (including sensation of bloating, early satiety, postprandial fullness, upper abdominal pain or discomfort [epigastric pain, epigastric burning], anorexia [loss of appetite], heartburn, nausea, and vomiting) after 4 and 8 weeks of treatment and responder analysis for adequate/satisfactory relief as assessed by the LDQ and/or NRS-11. Treatment acceptance and ease of use assessed by participants using the 5-point Likert scale was an exploratory endpoint. Safety and tolerability in both treatment arms were also evaluated, and endpoints included the overall summary of treatment-emergent adverse events (TEAEs), prolactin levels and change from baseline in prolactin. The primary and secondary efficacy endpoints were analyzed across subgroups (gender, age group, and geographical region) using the same analysis as specified for the primary, secondary, and safety endpoints.

### Statistical analyses

The design of this non-inferiority study included statistical considerations for sample size and non-inferiority margin. Based on previous studies, assuming an NI-margin of 1.75 and a standard deviation of 7 for the change in the LDQ severity score, 253 subjects per treatment arm were required to conclude non-inferiority between the two treatment groups with a power of 80% at an α-level of 2.5% single-sided. On adding a 10% drop-out rate, the total sample size was calculated as 564 subjects (N=282 per arm). The change in LDQ severity and NRS-11 symptom scores were analyzed using the analysis of covariance (ANCOVA) model. All secondary efficacy endpoints were assessed using descriptive statistics including frequency count and percentages by group, as were adverse events, laboratory values, and vital signs.

## Results

Of 564 enrolled participants, 96.3% (97.2% in the TID group and 95.4% in the OD group) completed the study, and 21 (3.7%) participants prematurely discontinued the study.

The total number of participants included in the safety population, defined as all participants who had received at least one dose of Itopride hydrochloride, was 561 (281 and 280 participants in the TID and OD groups, respectively). The full analysis (FA) population, defined as all participants in the safety population who had at least one post-baseline assessment of the efficacy parameters, totaled 557 (including 280 and 277 participants in the TID and OD groups, respectively). The per protocol (PP) population, comprised of all participants in the FA population who had no significant protocol deviations, totaled 524 (including 263 and 261 participants in the TID and OD groups, respectively).

The mean age of participants in both treatment groups was 40.32 (standard deviation [SD] 13.93) years, with the age of the majority of participants ranging from 18 to 44 years. The two treatment groups were comparable for age distribution, gender, ethnicity, baseline characteristics, medical history, use of concomitant medications, and treatment compliance (≥95% in both groups; **Table 1**).

**Table 1.** Demographics and baseline characteristics of all participants.

|  | Statistics | Itopride hydrochloride 50 mg TID<br>(n = 282) | Itopride hydrochloride 150 mg OD<br>(n = 282) | All participants<br>(N = 564) |
| --- | --- | --- | --- | --- |
| **Age (years)** | Mean (SD);<br>range | 40.86 (13.71); 18–79 | 39.78 (14.15); 18–77 | 40.32 (13.93); 18–79 |
| **Gender** |  |  |  |  |
| Male | n (%) | 90 (31.9) | 67 (23.8) | 157 (27.8) |
| Female | n (%) | 192 (68.1) | 215 (76.2) | 407 (72.2) |
| **Race** |  |  |  |  |
| White | n (%) | 104 (36.9) | 107 (37.9) | 211 (37.4) |
| Asian | n (%) | 178 (63.1) | 175 (62.1) | 353 (62.6) |
| **Body Mass Index (kg/m<sup>2</sup>)** | Mean (SD);<br>range | 25.67 (4.65); 16.4–40.4 | 25.56 (4.78); 15.1–43.7 | 25.62 (4.71); 15.1–43.7 |
| **At Least One Medical History Finding\*** | n (%) | 113 (40.1) | 113 (40.1) | 226 (40.1) |
| **Concomitant Medications** |  |  |  |  |
| Any Concomitant Medication | n (%) | 90 (31.9) | 95 (33.7) | 185 (32.8) |
| Lipid Modifying Agents, Plain (C10A) | n (%) | 31 (11.0) | 24 (8.5) | 55 (9.8) |
| Selective Calcium Channel Blockers with Mainly Vascular Effects (C08C) | n (%) | 17 (6.0) | 21 (7.4) | 38 (6.7) |
| ACE Inhibitors, Plain (C09A) | n (%) | 15 (5.3) | 14 (5.0) | 29 (5.1) |
| Beta-Blocking Agents (C07A) | n (%) | 14 (5.0) | 14 (5.0) | 28 (5.0) |
| Other Analgesics and Antipyretics (N02B) | n (%) | 15 (5.3) | 13 (4.6) | 28 (5.0) |
| **Overall Study Drug Compliance** | n, Mean (SD); range | 281, 94.34 (7.49); 33.3–104.8 | 277, 97.97 (10.67); 55.0–242.9 | 558, 96.14 (9.38); 33.3–242.9 |

\*Medical history findings reported by 5 or more subjects, including menopause, post-menopause, hypertension, dyslipidemia, diabetes mellitus, type 2 diabetes mellitus, obesity, asthma, rhinitis allergic, osteoarthritis, hypothyroidism, hyperthyroidism, drug hypersensitivity and migraine.
OD, once-daily; SD, standard deviation; TID, three times a day.

### Efficacy

In the PP population, change in LDQ severity score after 8 weeks showed comparable efficacy between the two treatment groups with a least-squares (LS) mean reduction of –9.17 (95% CI –9.78, –8.56) and –9.54 (95% CI –10.17, –8.91) from baseline to Week 8 for the TID and OD groups, respectively (**Figure 2A**, which shows the mean change in LDQ score from baseline to Week 8). The results also demonstrated that itopride hydrochloride 150 mg OD is non-inferior to itopride hydrochloride 50 mg TID, with the lower bounds of the 95% CI for the LS mean difference between treatment groups (–0.21), exceeding the prespecified non-inferiority margin of –1.75. The FA population confirmed the findings with an LS mean change in LDQ severity score from baseline to Week 8 of –9.60 (95% CI –10.15, –9.05) in the TID group and –9.76 (95% CI –10.32, –9.19) in the OD group. These findings also showed that itopride hydrochloride 150 mg OD is non-inferior to itopride hydrochloride 50 mg TID, with the lower bounds of the 95% CI for the LS mean difference between treatment groups (–0.42) exceeding the prespecified non-inferiority margin of –1.75, demonstrating the comparable efficacy of the two treatment regimens (**Figure 2B**, which shows the mean change in LDQ score from baseline to Week 8).

**Figure 2.**
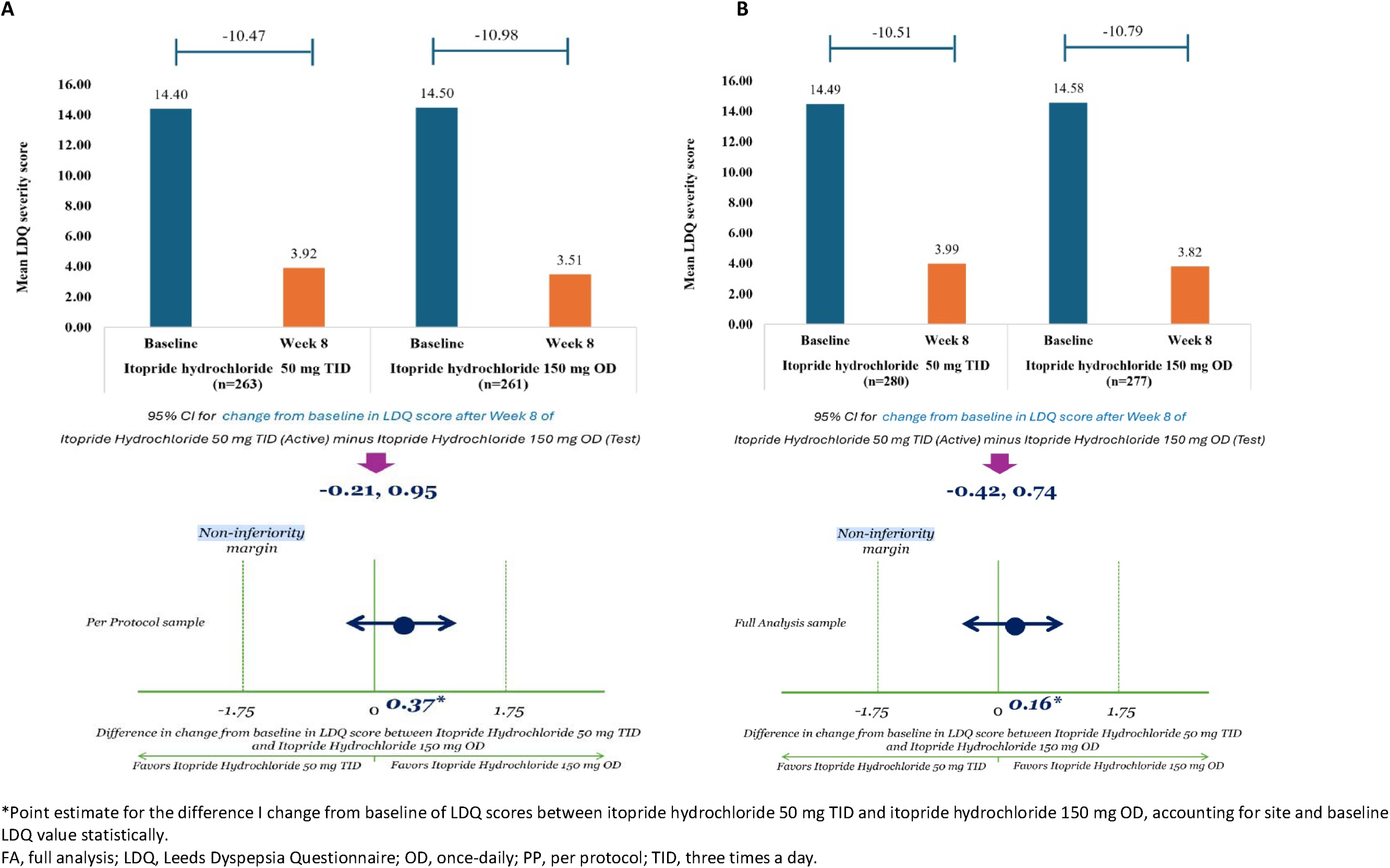
Change in mean LDQ severity score after 8 weeks of treatment from baseline in the (A) PP population and (B) FA population

In the FA population, the mean change in LDQ severity score was also examined from baseline to Week 4, showing that itopride hydrochloride 50 mg TID and itopride hydrochloride 150 mg OD were comparable with a treatment difference of 0.21 [95% CI –0.44, 0.86], thus indicating that extended release OD itopride hydrochloride was non-inferior to immediate release itopride hydrochloride (**Figure 3**).

**Figure 3.**
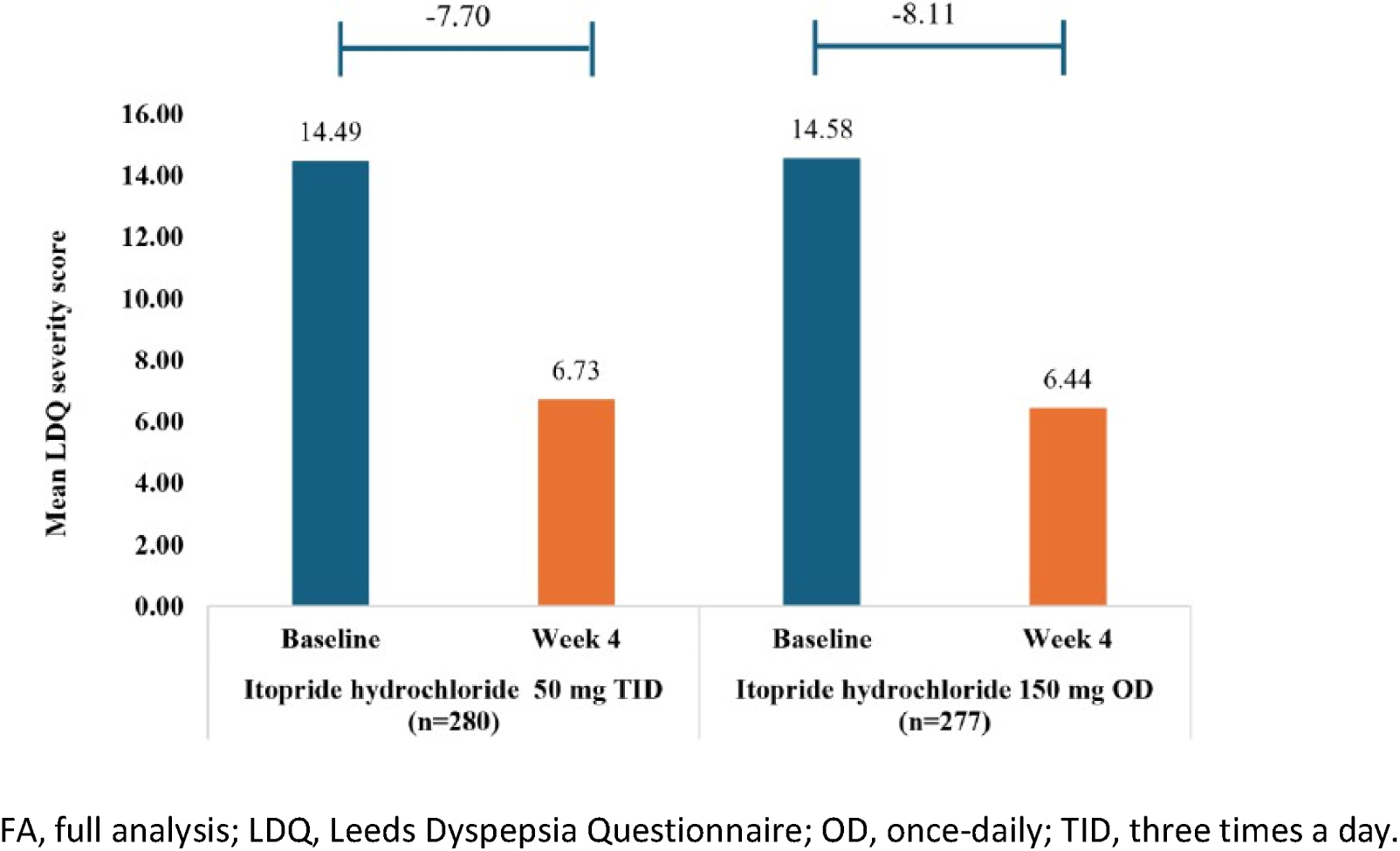
Change in mean LDQ severity score after 4 weeks of treatment from baseline in the FA population

The SF-NDI questionnaire showed comparable improvements in FD scores from baseline to Week 8 in both treatment groups (**Supplementary Table 1**). The mean overall QoL transformed scores were similar between the two treatment groups at baseline (p=0.856) and Week 8 (p=0.985; **Table 2**). No notable differences were observed between groups in any of the five subscale scores (tension, interference with daily activities, eating/drinking, knowledge/control, work/study).

**Table 2.** Transformed QoL Scores assessed by the SF-NDI Questionnaire in the FA population.

| Characteristics | Statistics | Baseline |  | Week 8 |  |
| --- | --- | --- | --- | --- | --- |
|  |  | Itopride hydrochloride | Itopride hydrochloride | Itopride hydrochloride | Itopride hydrochloride |
|  |  | 50 mg TID (n = 280) | 150 mg OD (n = 277) | 50 mg TID (n = 278) | 150 mg OD (n = 274) |
| Overall QoL Transformed Score |  |  |  |  |  |
|  | Mean (SD) | 67.96 (19.651) | 68.27 (19.795) | 90.29 (14.096) | 90.26 (14.281) |
|  | p value |  |  | 0.985 |  |
| Tension Domain |  |  |  |  |  |
|  | Mean (SD) | 64.73 (23.200) | 65.21 (22.459) | 88.08 (16.834) | 88.91 (16.601) |
|  | p value |  |  | 0.560 |  |
| Interference with Daily Activities Domain |  |  |  |  |  |
|  | Mean (SD) | 71.79 (23.659) | 71.44 (22.902) | 92.54 (14.386) | 91.88 (17.024) |
|  | p value |  |  | 0.625 |  |
| Eating/Drinking Domain |  |  |  |  |  |
|  | Mean (SD) | 65.40 (24.295) | 64.26 (24.583) | 87.32 (17.740) | 87.41 (16.436) |
|  | p value |  |  | 0.951 |  |

| Knowledge/Control Domain |  |  |  |  |  |
| --- | --- | --- | --- | --- | --- |
|  | Mean (SD) | 66.65 (23.309) | 65.93 (25.881) | 90.92 (15.267) | 89.78 (16.241) |
|  | p value |  |  |  | 0.397 |
| Work/Study Domain |  |  |  |  |  |
|  | Mean (SD) | 71.25 (26.297) | 74.50 (25.119) | 92.58 (16.127) | 93.34 (15.728) |
|  | p value |  |  |  | 0.576 |

FA, full analysis; QoL, quality of life; SD, standard deviation; SF-NDI, Short Form Nepean Dyspepsia Index.

Analysis of the NRS-11 scores showed marked improvement in the severity of all FD symptoms in both the OD and TID treatment groups after 4 and 8 weeks of treatment, with comparable improvement in the two groups (**Supplementary Table 2**).

The responder rates (based on LDQ and NRS-11 scores) were similar in both treatment groups after 8 weeks of treatment, with an 84.6% and 86.3% response rate in the TID and OD groups, respectively.

After 8 weeks of treatment, the number of participants responding as “very satisfied” and “extremely satisfied” with the dosing schedule was higher in the OD group compared with the TID group (**Table 3**). Additionally, the mean treatment acceptance score was significantly higher in the OD group versus the TID group (4.22 vs. 3.83; p<0.001), indicating greater acceptance and preference for the OD dosing regimen.

**Table 3.** Treatment acceptance and ease of use in the FA population.

|  | Statistics | Itopride hydrochloride<br>50 mg TID (n = 280) | Itopride hydrochloride<br>150 mg OD (n = 277) |
| --- | --- | --- | --- |
| **Response on 5-point Likert Scale** |  |  |  |
| Missing | n (%) | 2 (0.7) | 3 (1.1) |
| 1 – Not at all satisfied | n (%) | 9 (3.2) | 1 (0.4) |
| 2 – Slightly satisfied | n (%) | 23 (8.2) | 8 (2.9) |
| 3 – Neutral | n (%) | 55 (19.6) | 32 (11.6) |
| 4 – Very satisfied | n (%) | 109 (38.9) | 122 (44.0) |
| 5 – Extremely satisfied | n (%) | 82 (29.3) | 111 (40.1) |
| **Treatment Acceptance** |  |  |  |
|  | n | 278 | 274 |
|  | Mean (SD) | 3.83 (1.05) | 4.22 (0.79) |
|  | p value | < 0.001 |  |

FA, full analysis; OD, once-daily; SD, standard deviation; TID, three times a day.

Subgroup analyses by gender, age group, and geographical region were consistent across all primary, secondary, and exploratory endpoints (**Supplementary Table 3**).

### Safety

The majority of AEs were mild or moderate in intensity in both treatment arms. No clinically significant impact of both dosing regimens was observed on laboratory parameters, weight, or vital signs. The overall incidence of TEAEs was slightly higher in the OD group (26.1%; 110 TEAEs) than in the TID group (17.4%; 64 TEAEs). The most frequently occurring individual TEAE (occurring in ≥3% of subjects) was diarrhea, which was reported more frequently with itopride hydrochloride 150 mg OD (5.7%) than with itopride hydrochloride 50 mg TID group (3.6%; **Supplementary Table 4**).

As assessed by the Investigator, the incidence of TEAEs that were considered to have at least a reasonable possibility of a causal relationship to the study drug was higher in the OD group (13.2%; 51 TEAEs) compared with the TID group (7.5%; 24 TEAEs). The most common TEAEs that had a reasonable possibility for a causal relationship in the TID and OD groups, respectively, were diarrhea (8 [2.8%] vs 13 [4.6%]), dizziness (3 [1.1%] vs 7 [2.5%]), nausea (2 [0.7%] vs 5 [1.8%]), and headache (4 [1.4%] in both groups). Treatment-related TEAEs across both arms were predominantly mild and transient. No new or unexpected safety signals emerged, and the pattern of related events was consistent with the established safety profile of itopride hydrochloride. Hyperprolactinemia was reported with reasonable relationship in four participants (three in the TID group and one in the OD group), consistent with the known dopamine D2 receptor antagonism of itopride hydrochloride; these findings were not deemed clinically significant. Overall, the mean change in prolactin levels remained stable over the 8-week treatment period in both treatment groups

TEAEs that resulted in study termination were reported in 0.7% (3 TEAEs) and 2.1% (9 TEAEs) of participants in the TID and OD groups, respectively, and the outcome of all TEAEs was recovered or resolved.

Severe TEAEs were observed in 0.7% (2 TEAEs) of the participants in the TID group, including abdominal pain (reported as a TEAE) and pulmonary embolism (reported as a treatment-emergent serious adverse event [TESAE]), compared with 1.1% (4 TEAEs) of participants in the OD group (including diarrhea, upper abdominal pain, headache, and dyspnea). No deaths were reported during the study (**Supplementary Table 4**).

No TESAEs were reported in the OD group, while two TESAEs were reported in 2 (0.7%) participants in the TID group: urinary tract infection (0.4%; moderate severity) and pulmonary embolism (0.4%; severe). Both TESAEs were assessed as unrelated to the study drug.

## Discussion

This study revealed that an OD 150 mg extended-release tablet of itopride hydrochloride is non-inferior to thrice-daily 50 mg immediate-release tablets and has proven efficacy for managing FD. Both products showed substantial improvement in LDQ severity scores, control of dyspepsia symptoms, enhanced disease-specific QoL, and alleviation of unpleasant symptoms associated with gastric dysmotility and delayed gastric emptying. The majority of participants reported satisfactory or adequate relief following either TID or OD treatment across the LDQ and NRS-11 scales. In elderly patients, there was a marked improvement in LDQ severity scores with the OD 150 mg extended-release tablet, potentially reflecting the benefit of a simplified dosing schedule—particularly for older adults who may already be managing multiple medications.

Supportive pharmacokinetic data from Phase 1 further contextualize these clinical findings. Prior to Phase 3, a single-dose study in 36 healthy adults assessed the effect of food on the bioavailability of the OD 150 mg extended-release formulation under fasting, high-fat fed, and normocaloric meal conditions (**Supplementary Figure 1**). The OD tablet demonstrated predictable absorption characteristics across all conditions, including the proposed label administration (30 minutes before a normocaloric meal), with no unexpected release pattern observed. The study also compared exposure between the OD 150 mg and TID 50 mg formulations and showed comparable area under the curve (AUC) values (**Supplementary Table 5**). A complementary multiple-dose, 5-day crossover study in 30 healthy subjects confirmed steady-state bioequivalence between the OD and TID regimens, with the geometric mean ratio for AUC(0-τ)_ss within the accepted 80–125% range (**Supplementary Tables 6 and 7**; **Supplementary Figure 2**). Together, these Phase 1 findings demonstrate that the extended-release formulation provides consistent drug exposure, supporting its suitability as an OD alternative and reinforcing the validity of the Phase 3 clinical comparisons.

The itopride hydrochloride 150 mg OD tablets also showed better acceptance and ease of use, offering a more convenient dosing regimen that could improve compliance and treatment outcomes in real-world practice. The significantly higher mean treatment-acceptance score in the OD group (4.22 vs 3.83; p<0.001) aligns with previous evidence demonstrating that reducing dosing frequency improves adherence across both acute and chronic therapeutic settings (18–20). Moreover, while consistent daily timing is recommended, the OD regimen allowed flexibility to align intake with individual eating patterns—such as taking the dose with lunch rather than breakfast—which may further contribute to patient satisfaction.

Both dosing regimens were well tolerated, with TEAEs mostly mild and predominantly gastrointestinal, consistent with the known safety profile of itopride hydrochloride. The slightly higher frequency of TEAEs in the OD group did not translate into clinically significant safety concerns, and no meaningful differences were observed in laboratory parameters, vital signs, or weight. Overall, these findings support a positive benefit–risk profile for the OD 150 mg extended-release formulation.

FD presents subjective and often overlapping symptoms, making multidimensional assessment essential. The secondary endpoints, including SF-NDI and NRS-11 symptom scores, confirmed consistent improvements across both groups. The SF-NDI results aligned with previous validation work in Asian populations (21) with participants moving from severe dyspepsia at baseline to mild symptom burden by Week 8. This mirrors the improvement observed across individual symptoms and suggests that symptom relief translated into enhanced daily functioning and overall QoL.

Itopride hydrochloride acts by enhancing gastrointestinal motility and accelerating gastric emptying, thereby addressing the underlying pathophysiological mechanisms of gastric dysmotility and delayed gastric emptying, which are common across FD subtypes. Clinically, itopride improves both meal-related symptoms (e.g. bloating, early satiety, postprandial fullness) and upper abdominal pain or discomfort, regardless of subtype classification covering the efficacy and safety for epigastric pain syndrome (EPS), postprandial distress syndrome (PDS), or overlapping PDS and EPS. This is consistent with recent clinical evidence from a randomized controlled trial showing that formal differentiation between EPS and PDS did not yield superior outcomes over empirical therapy, with comparable global response rates at 8 weeks (74.4% vs 72.2%) and similar improvements in individual symptoms and quality of life across subtypes (22). The authors attributed these findings to the substantial mechanistic overlap between EPS and PDS, including disordered gastric accommodation, visceral hypersensitivity, duodenal inflammation, and reflux physiology. This supports the clinical relevance of prokinetic therapy such as itopride across the broader FD population rather than restricted to formally defined PDS.

Strengths of this study include a large, ethnically diverse patient population spanning a wide age range, ensuring generalizability. Baseline characteristics were well balanced across groups, indicating minimal confounding. The open-label design, while sometimes considered a limitation, was an important strength in assessing real-world treatment acceptance and ease of use, which are key attributes for an OD regimen. Other strengths include the selected evaluations. Efficacy in patients diagnosed with functional dyspepsia according to Rome IV is evaluated using validated patient-reported outcome instruments that quantify symptom severity and frequency. The LDQ, including its validated short-form versions, is a well-established instrument developed to measure the burden of dyspeptic symptoms. It generates a quantitative score reflecting the frequency and severity of key dyspepsia symptoms and has demonstrated reliability, construct validity, and responsiveness to change in clinical studies. Multiple studies have used LDQ for FD as a primary endpoint and it was used as the reference for this Phase 3 study in the sample size calculation. The Rome IV criteria are applied to establish the diagnosis of functional dyspepsia, while the LDQ is used to assess baseline symptom severity and to evaluate changes in symptoms over time. The combined use of Rome IV for diagnostic classification and LDQ for efficacy assessment represents a complementary and methodologically appropriate approach, supporting the demonstration of clinically meaningful treatment effects in patients with functional dyspepsia. A limitation is the absence of site-specific analyses despite the study being conducted across 19 sites in 5 countries. Meanwhile, future work could explore regional variability in treatment response.

The clinical implications of an OD 150 mg extended-release formulation are substantial. By reducing pill burden and offering a more convenient dosing schedule without compromising efficacy or safety, the OD regimen supports better adherence and may translate into improved long-term management of FD (and chronic gastritis). This aligns well with patient preferences and contemporary therapeutic goals centered on simplifying treatment while maintaining clinical benefit.

## Conclusions

The study confirmed the sustained release profile of itopride over a prolonged period of time without signs of unexpected release characteristics like dose-dumping and demonstrated that the exposure of the OD 150 mg extended-release tablet was bioequivalent to the TID 50 mg immediate-release tablets. This Phase 3 clinical study directly comparing the extended-release and immediate-release forms of itopride hydrochloride showed the OD 150 mg extended-release tablet was non-inferior to thrice-daily 50 mg immediate-release tablets with comparable efficacy for managing FD (and chronic gastritis). Moreover, the OD 150 mg regimen could potentially be of greater therapeutic benefit in elderly patients compared with the thrice-daily 50 mg regimen. The steady state study also showed that both OD 150 mg tablets and 50 mg tablets TID were bioequivalent in terms of extent of exposure at steady state.

Both dosing regimens were well-tolerated when administered to healthy subjects as well as patients, with overall assessments suggesting a positive benefit-risk profile of the OD 150 mg extended-release itopride hydrochloride formulation.

## Supporting information

ITOPRIDE in FD MS_Final Version 12July2026 supplement

## Data Availability

The data that support the findings of this study are available from the corresponding author, upon reasonable request.

## Statement of authorship

Authors equally contributed equally to the conception, design and writing of the manuscript. All authors critically revised the manuscript, agree to be fully accountable for ensuring the integrity and accuracy of the work, and read and approved the final manuscript.

## Acknowledgments

Editorial assistance was provided by Martin Guppy at Metamols Limited.

