## Supplementary material for "Itopride hydrochloride extended-release vs film-coated tablets for gastrointestinal symptoms in functional dyspepsia": ITOPRIDE in FD MS_Final Version 12July2026 supplement

### Supplementary data

**Supplementary Table 1.** Summary of Symptoms and Functional Dyspepsia Score assessed by SF-NDI Questionnaire in the FA population

| **Symptom score** | **Itopride hydrochloride 50 mg TID (n = 280)** | | **Itopride hydrochloride 150 mg OD (n = 277)** | |
| --- | --- | --- | --- | --- |
|  | **Baseline mean (SD); range** | **Week 8 mean (SD); range** | **Baseline mean (SD); range** | **Week 8 mean (SD); range** |
| Pain or ache in upper abdomen | 4.57 (3.02); 0–12 | 1.42 (2.30); 0–12 | 4.35 (3.20); 0–13 | 1.43 (2.50); 0–12 |
| Discomfort in upper abdomen | 4.86 (2.95); 0–12 | 1.51 (2.45); 0–12 | 4.49 (3.15); 0–13 | 1.49 (2.42); 0–12 |
| Burning sensation in upper  abdomen | 3.93 (3.04); 0–13 | 1.34 (2.24); 0–11 | 3.97 (3.26); 0–13 | 1.35 (2.30); 0–12 |
| Burning sensation in chest  (heartburn) | 3.78 (3.09); 0–13 | 1.31 (2.23); 0–12 | 3.56 (3.18); 0–13 | 1.30 (2.15); 0–11 |
| Cramps in upper abdomen | 1.59 (2.66); 0–11 | 0.38 (1.40); 0–9 | 1.88 (2.85); 0–13 | 0.45 (1.60); 0–10 |
| Pain or ache in chest | 1.58 (2.63); 0–12 | 0.43 (1.55); 0–12 | 1.71 (2.80); 0–13 | 0.54 (1.89); 0–12 |
| Inability to finish a regular meal | 2.71 (3.32); 0–13 | 0.65 (2.09); 0–13 | 2.73 (3.37); 0–13 | 0.81 (2.17); 0–13 |
| Bitter/sour tasting fluid that comes up into your mouth or  throat | 3.93 (3.18); 0–13 | 1.10 (2.20); 0–13 | 3.55 (3.02); 0–13 | 1.04 (1.94); 0–12 |
| Fullness after eating or slow  digestion | 5.44 (3.36); 0–13 | 1.92 (2.75); 0–13 | 5.49 (3.30); 0–13 | 2.06 (2.82); 0–13 |
| Pressure in upper abdomen | 2.96 (3.27); 0–12 | 0.89 (2.06); 0–13 | 3.32 (3.46); 0–13 | 0.93 (2.13); 0–13 |
| Bloating in upper abdomen | 4.81 (3.29); 0–12 | 1.62 (2.46); 0–13 | 4.86 (3.50); 0–13 | 1.97 (2.78); 0–13 |

| Nausea | 2.93 (3.00); 0–12 | 0.96 (2.02); 0–13 | 2.60 (3.00); 0–13 | 0.95 (2.06); 0–11 |
| --- | --- | --- | --- | --- |
| Burping/belching | 4.29 (3.60); 0–13 | 1.56 (2.68); 0–13 | 4.65 (3.72); 0–13 | 1.31 (2.55); 0–13 |
| Vomiting | 1.18 (2.48); 0–13 | 0.28 (1.29); 0–10 | 0.92 (2.21); 0–13 | 0.19 (0.88); 0–6 |
| Bad breath | 2.09 (3.05); 0–12 | 0.85 (1.93); 0–11 | 1.83 (2.95); 0–12 | 0.76 (1.89); 0–11 |
| Functional dyspepsia score* | 16.65 (8.65); 0–50 | 5.33 (7.55); 0–46 | 16.55 (9.28); 0–49 | 5.65 (7.87); 0–47 |

*Functional dyspepsia score was calculated as the sum of 4 symptoms: pain or ache in upper abdomen, burning sensation in upper abdomen, inability to finish a regular meal and fullness after eating or slow digestion.

FA, full analysis; OD, once-daily; SD, standard deviation; SF-NDI, Short Form Nepean Dyspepsia Index; TID, three times a day.

**Supplementary Table 2.** Change in NRS-11 Score for Symptoms after Week 4 and Week 8 in the FA population

| **Symptom** | **Statistics** | **Itopride hydrochloride 50 mg TID (n = 280)** | | | | | **Itopride hydrochloride 150 mg OD (n = 277)** | | | | |
| --- | --- | --- | --- | --- | --- | --- | --- | --- | --- | --- | --- |
|  |  | **Baseline** | **Week 4** | **Change from baseline to Week 4** | **Week 8** | **Change from baseline to Week 8** | **Baseline** | **Week 4** | **Change from baseline to Week 4** | **Week 8** | **Change from baseline to Week 8** |
| **Bloating sensation** | Mean (SD) | 1.91  (1.92) | 0.46  (1.08) | -1.43 (1.70) | 0.37  (0.95) | -1.54 (1.81) | 2.00  (2.102) | 0.57  (1.21) | -1.41  (2.01) | 0.35  (0.97) | -1.63  (2.00) |
|  | LS mean difference* (SE); 95% CI |  |  | -0.08 (0.09);  -0.25, 0.09 |  | 0.04 (0.08);  -0.11, 0.18 |  |  |  |  |  |
| **Early satiety** | Mean (SD) | 1.83  (2.05) | 0.43  (1.14) | -1.38 (1.87) | 0.33  (0.91) | -1.50 (1.88) | 1.74  (2.010) | 0.53  (1.22) | -1.20  (1.90) | 0.34  (0.99) | -1.40  (1.89) |
|  | LS mean difference* (SE); 95% CI |  |  | -0.10 (0.09);  -0.28, 0.08 |  | -0.01 (0.07);  -0.16, 0.13 |  |  |  |  |  |
| **Postprandial fullness** | Mean (SD) | 2.35  (2.18) | 0.44  (1.13) | -1.90 (1.94) | 0.36  (0.92) | -2.00 (1.97) | 2.21  (2.155) | 0.58  (1.19) | -1.61  (2.10) | 0.38  (1.02) | -1.81  (1.99) |
|  | LS mean difference* (SE); 95% CI |  |  | -0.15 (0.09);  -0.33, 0.02 |  | -0.05 (0.07);  -0.19, 0.10 |  |  |  |  |  |
|  | Mean (SD) | 2.17  (2.08) | 0.34  (0.80) | -1.81 (1.92) | 0.32  (0.89) | 0.32 (0.99) | 1.96  (2.270) | 0.48  (1.16) | -1.46  (2.12) | 0.32  (0.99) | -1.63  (2.12) |

| **Upper abdominal pain or discomfort** | LS mean difference* (SE); 95% CI |  |  | -0.15  (0.073);  -0.29,  -0.00 |  | -0.03  (0.072);  -0.17, 0.12 |  |  |  |  |  |
| --- | --- | --- | --- | --- | --- | --- | --- | --- | --- | --- | --- |
| **Anorexia (loss of appetite)** | Mean (SD) | 0.90  (1.76) | 0.16  (0.63) | -0.72 (1.69) | 0.14  (0.76) | -0.76 (1.57) | 1.05  (1.88) | 0.24  (0.79) | -0.78  (1.57) | 0.19  (0.88) | -0.83  (1.66) |
|  | LS mean difference* (SE); 95% CI |  |  | -0.04 (0.05);  -0.14, 0.07 |  | 0.00 (0.06);  -0.12, 0.13 |  |  |  |  |  |
| **Heartburn** | Mean (SD) | 1.65  (1.98) | 0.24  (0.63) | -1.41 (1.88) | 0.20  (0.74) | -1.44 (1.91) | 1.64  (1.92) | 0.32  (0.95) | -1.29  (1.81) | 0.22  (0.76) | -1.39  (1.73) |
|  | LS mean difference* (SE); 95% CI |  |  | -0.09 (0.06);  -0.21, 0.03 |  | -0.01 (0.06);  -0.13, 0.10 |  |  |  |  |  |
| **Nausea** | Mean (SD) | 1.01  (1.60) | 0.14  (0.48) | -0.84 (1.54) | 0.13  (0.59) | -0.87 (1.57) | 0.85  (1.53) | 0.17  (0.59) | -0.69  (1.49) | 0.15  (0.58) | -0.71  (1.45) |
|  | LS mean difference* (SE); 95% CI |  |  | -0.02 (0.04);  -0.11, 0.06 |  | -0.03 (0.05);  -0.13, 0.06 |  |  |  |  |  |
| **Vomiting** | Mean (SD) | 0.34  (1.22) | 0.07  (0.66) | -0.27 (1.03) | 0.03  (0.29) | -0.31 (1.15) | 0.31  (1.08) | 0.07  (0.39) | -0.25  (1.13) | 0.03  (0.30) | -0.30  (1.14) |
|  | LS mean difference* (SE); 95% CI |  |  | 0.00 (0.04);  -0.08, 0.09 |  | 0.01 (0.03);  -0.04, 0.06 |  |  |  |  |  |

*LS mean difference is calculated comparing the two treatment groups (50 mg TID versus 150 mg OD).

CI, confidence interval; FA, full analysis; LS, least squares; NRS-11, Numerical Rating Scale – 11; OD, once-daily; SD, standard deviation; SE, standard error; TID, three times a day.

**Supplementary Table 3.** Subgroup analyses by gender, age group, and geographical region of change in LDQ severity score in the FA population

| **Subgroup** | **Statistics** | **Itopride hydrochloride 50 mg TID (n = 280)** | | | | | **Itopride hydrochloride 150 mg OD (n = 277)** | | | | |
| --- | --- | --- | --- | --- | --- | --- | --- | --- | --- | --- | --- |
|  |  | **Baseline** | **Week 4** | **Change from baseline to Week 4** | **Week 8** | **Change from baseline to Week 8** | **Baseline** | **Week 4** | **Change from baseline to Week 4** | **Week 8** | **Change from baseline to Week 8** |
| **Male** | Mean (SD) | 14.46  (4.37) | 5.92  (4.33) | -8.52 (5.12) | 3.20  (3.907) | -11.26 (4.83) | 14.58  (4.12) | 6.98  (5.09) | -7.59  (4.40) | 4.23  (5.08) | -10.40  (4.59) |
|  | LS mean difference* (SE); 95% CI |  |  | -1.18  (0.636);  -2.44, 0.08; p=0.066 |  | -0.94 (0.57);  -2.07, 0.19; p=0.103 |  |  |  |  |  |
| **Female** | Mean (SD) | 14.50  (3.98) | 7.11  (5.23) | -7.32 (5.46) | 4.37  (4.649) | -10.15 (4.69) | 14.59  (4.78) | 6.27  (4.28) | -8.27  (5.08) | 3.69  (3.89) | -10.91  (5.12) |
|  | LS mean difference* (SE); 95% CI |  |  | 0.69 (0.39); -  0.08, 1.46; p=0.078 |  | 0.65 (0.35); -  0.04, 1.33; p=0.063 |  |  |  |  |  |
| **18**–**44 years of age** | Mean (SD) | 14.93  (4.21) | 6.82  (5.13) | -8.03 (5.47) | 4.02  (4.568) | -10.91 (4.91) | 14.83  (4.82) | 6.68  (4.67) | -8.14  (5.07) | 4.08  (4.63) | -10.80  (5.23) |
|  | LS mean difference* (SE); 95% CI |  |  | -0.08 (0.42);  -0.90, 0.74; p=0.850 |  | -0.09 (0.39);  -0.86, 0.67; p=0.812 |  |  |  |  |  |
| **45–59 years of age** | Mean (SD) | 13.97  (3.94) | 6.19  (4.43) | -7.79 (5.51) | 3.49  (4.01) | -10.47 (4.78) | 14.34  (4.38) | 5.98  (3.83) | -8.22  (4.62) | 3.19  (3.12) | -11.15  (4.66) |

|  | LS mean difference* (SE); 95% CI |  |  | 0.09 (0.71); -  1.33, 1.50; p=0.905 |  | -0.09  (0.536);  -1.16, 0.97; p=0.860 |  |  |  |  |  |
| --- | --- | --- | --- | --- | --- | --- | --- | --- | --- | --- | --- |
| **≥60 years of age** | Mean (SD) | 13.09  (3.39) | 7.39  (5.37) | -5.65 (4.28) | 4.90  (4.650) | -8.26 (3.055) | 13.68  (3.54) | 5.94  (4.55) | -7.74  (4.76) | 3.52  (3.23) | -10.09  (4.23) |
|  | LS mean difference* (SE); 95% CI |  |  | 1.99 (0.98);  0.03, 3.94; p=0.047 |  | 2.26 (0.706);  0.84, 3.68; p=0.002 |  |  |  |  |  |
| **Armenia** | Mean (SD) | 13.38  (3.88) | 7.25  (3.80) | -5.99 (3.60) | 4.32  (4.257) | -9.07 (3.949) | 13.14  (3.88) | 6.89  (3.73) | -6.26  (3.69) | 4.09  (3.41) | -9.08  (3.77) |
|  | LS mean difference* (SE); 95% CI |  |  | 0.22 (0.442);  -0.65, 1.10; p=0.612 |  | 0.21 (0.448);  -0.68, 1.09; p=0.646 |  |  |  |  |  |
| **Philippines** | Mean (SD) | 14.51  (3.90) | 4.23  (4.40) | -10.28 (5.31) | 1.48  (2.539) | -13.02  (4.326) | 14.70  (4.21) | 3.92  (3.34) | -10.77  (4.53) | 1.34  (2.43) | -13.36  (4.08) |
|  | LS mean difference* (SE); 95% CI |  |  | 0.17 (0.50); -  0.83, 1.16; p=0.742 |  | 0.03 (0.32); -  0.61, 0.67; p=0.926 |  |  |  |  |  |
| **Malaysia** | Mean (SD) | 15.34  (3.65) | 9.84  (4.85) | -5.58 (5.32) | 7.29  (4.414) | -8.19 (4.85) | 17.97  (5.54) | 10.75  (6.07) | -7.11  (6.97) | 8.63  (6.34) | -9.33  (7.55) |
|  | LS mean difference* (SE); 95% CI |  |  | 0.06 (1.43); -  2.81, 2.94; p=0.965 |  | -0.54 (1.45);  -3.46, 2.37; p=0.711 |  |  |  |  |  |

| **Thailand** | Mean (SD) | 16.62  (4.82) | 8.30  (6.08) | -8.36 (6.46) | 6.38  (5.49) | -10.24 (5.48) | 15.03  (5.15) | 7.76  (4.21) | -7.27  (4.93) | 5.06  (3.20) | -10.09  (5.32) |
| --- | --- | --- | --- | --- | --- | --- | --- | --- | --- | --- | --- |
|  | LS mean difference* (SE); 95% CI |  |  | -0.26 (1.21);  -2.68, 2.15; p=0.827 |  | 0.87 (1.04); -  1.21, 2.95; p=0.405 |  |  |  |  |  |
| **Vietnam** | Mean (SD) | 15.24  (3.95) | 8.63  (6.54) | -6.50 (7.25) | 4.94  (4.479) | -10.31 (4.33) | 16.06  (4.26) | 7.75  (4.24) | -8.38  (3.44) | 4.81  (4.19) | -11.25  (4.51) |
|  | LS mean difference* (SE); 95% CI |  |  | 1.49 (1.72); -  2.04, 5.02; p=0.394 |  | 0.14 (1.53); -  3.00, 3.27; p=0.930 |  |  |  |  |  |

*LS mean difference is calculated comparing the two treatment groups (50 mg TID versus 150 mg OD).

CI, confidence interval; FA, full analysis ; LDQ, Leeds Dyspepsia Questionnaire; OD, once-daily; SD, standard deviation; SE, standard error; TID, three times a day.

**Supplementary Table 4.** Incidence of TEAEs in the Safety Sample

| **Primary SOC**  **PT (incidence >1% in any group)** | **Statistics** | **Itopride hydrochloride 50 mg TID (n = 281)** | **Itopride hydrochloride 150 mg OD (n = 280)** |
| --- | --- | --- | --- |
| **At least one TEAE** | n (%) | 49 (17.4) | 73 (26.1) |
|  | Number of events | 64 | 110 |
| **Gastrointestinal disorders** | n (%) | 16 (5.7) | 32 (11.4) |
|  | Number of events | 17 | 45 |
| **Diarrhea** | n (%) | 10 (3.6) | 16 (5.7) |
|  | Number of events | 10 | 19 |
| **Abdominal pain upper** | n (%) | 1 (0.4) | 3 (1.1) |
|  | Number of events | 1 | 4 |
| **Abdominal pain** | n (%) | 0 | 3 (1.1) |
|  | Number of events | 0 | 4 |
| **Nausea** | n (%) | 2 (0.7) | 5 (1.8) |
|  | Number of events | 2 | 5 |
| **Infections and infestations** | n (%) | 13 (4.6) | 12 (4.3) |
|  | Number of events | 14 | 12 |
| **Nasopharyngitis** | n (%) | 3 (1.1) | 2 (0.7) |
|  | Number of events | 3 | 2 |
| **Influenza** | n (%) | 1 (0.4) | 3 (1.1) |
|  | Number of events | 1 | 3 |

| **Nervous system disorders** | n (%) | 10 (3.6) | 15 (5.4) |
| --- | --- | --- | --- |
|  | Number of events | 10 | 16 |
| **Headache** | n (%) | 7 (2.5) | 6 (2.1) |
|  | Number of events | 7 | 7 |
| **Dizziness** | n (%) | 3 (1.1) | 8 (2.9) |
|  | Number of events | 3 | 8 |
| **Respiratory, thoracic and mediastinal disorders** | n (%) | 4 (1.4) | 9 (3.2) |
|  | Number of events | 6 | 11 |
| **Dyspnea** | n (%) | 1 (0.4) | 3 (1.1) |
|  | Number of events | 1 | 3 |
| **Endocrine disorders** | n (%) | 3 (1.1) | 1 (0.4) |
|  | Number of events | 3 | 1 |
| **Hyperprolactinemia** | n (%) | 3 (1.1) | 1 (0.4) |
|  | Number of events | 3 | 1 |
| **Investigations** | n (%) | 1 (0.4) | 2 (0.7) |
|  | Number of events | 1 | 2 |
| **Vascular disorders** | n (%) | 0 (0) | 3 (1.1) |
|  | Number of events | 0 | 3 |
| **Hypertension** | n (%) | 0 | 3 (1.1) |
|  | Number of events | 0 | 3 |

n, number of participants; OD, once-daily; PT, preferred term; TEAE, treatment-emergent adverse event; SOC, system organ class, TID, three times a day.

**Supplementary Table 5.** Summary of pharmacokinetic parameters for food effect evaluation of itopride hydrochloride OD 150 mg after single dose administration

| **Treatment B (high-fat high-calorie fed condition) vs Treatment A (fasting condition)** | | | | | |
| --- | --- | --- | --- | --- | --- |
| **Parameter** | **N** | **Geometric least square mean (GLSM)** | | **Ratio %** | **90% CI** |
|  |  | **Treatment (B)** | **Treatment (A)** |  |  |
| Cmax (ng/mL) | 33 | 258.70 | 169.69 | 152.54 | 142.79 – 162.96 |
| AUC(0-t)  (hr.ng/mL) | 33 | 2285.91 | 1996.081 | 114.52 | 109.88 – 119.36 |
| AUC(0-inf) (hr.ng/mL) | 33 | 2362.96 | 2071.05 | 114.10 | 109.66 – 118.71 |
| **Treatment C (30 min before a normo-caloric meal) vs Treatment A (fasting condition)** | | | | | |
| **Parameter** | N | **Geometric least square mean (GLSM)** | | **Ratio %** | **90% CI** |
|  |  | **Treatment (C)** | **Treatment (A)** |  |  |
| Cmax (ng/mL) | 32 | 208.67 | 169.60 | 123.04 | 115.09 – 131.54 |
| AUC(0-t)  (hr.ng/mL) | 32 | 2074.23 | 1996.08 | 103.92 | 99.66 – 108.36 |
| AUC(0-inf) (hr.ng/mL) | 32 | 2153.19 | 2071.05 | 103.97 | 99.88 – 108.22 |

AUC, area under the curve; CV, coefficient of variation; GLSM, geometric least square mean.

**Supplementary Table 6.** Summary of bioequivalence parameters of itopride after single dose administration

| **Parameter** | **N** | **Geometric least square mean (GLSM)** | | **Ratio %** | **90% CI** | **Intra-subject CV (%)** |
| --- | --- | --- | --- | --- | --- | --- |
|  |  | **Test product (C)** | **Reference product (D)** |  |  |  |
| AUC(0-t)  (hr.ng/mL) | 32 | 2074.23 | 2311.75 | 89.73 | 86.01–93.60 | 10.23 |

AUC, area under the curve; CV, coefficient of variation; GLSM, geometric least square mean.

**Supplementary Table 7.** Summary of bioequivalence parameters of itopride at steady state

| **Parameter** | **N** | **Geometric least square mean (GLSM)** | | **Ratio %** | **90% CI** | **Intra-subject CV (%)** |
| --- | --- | --- | --- | --- | --- | --- |
|  |  | **Test product (A)** | **Reference product (B)** |  |  |  |
| AUC(0-τ)ss (hr.ng/mL) | 28 | 2188.27 | 2465.40 | 88.76 | 85.08, 92.60 | 9.29 |

CV, coefficient of variation; GLSM, geometric least square mean.

**Supplementary Figure 1.** Linear mean plasma concentration-time profiles of itopride after single dose administration of one extended release 150 mg tablet under fasted and fed conditions (Treatments A to C) and after TID dosing of a IR 50 mg tablet (Treatment D)

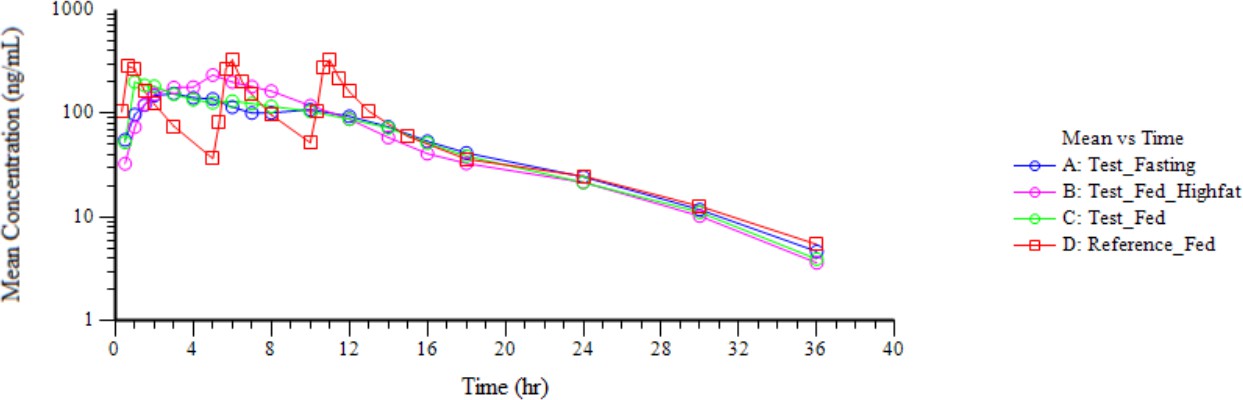

hr, hour.

**Supplementary Figure 2.** Linear mean graph of itopride hydrochloride at steady state (Day 5)

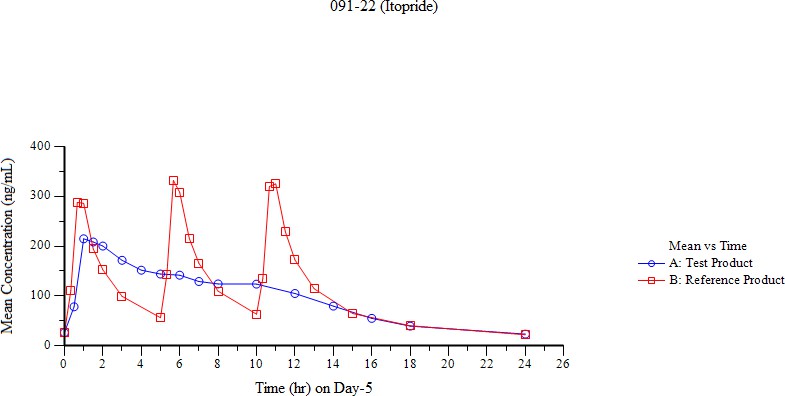

hr, hour.
